# The Associations of Diabetes Mellitus and Obesity on Osteoporosis

**DOI:** 10.64898/2026.03.16.26348517

**Authors:** Mendon G. Thomas, Ambalangodage C. Jayasuriya

## Abstract

**Objectives:** Osteoporosis is a common and debilitating condition that disproportionately affects older adults, particularly women, leading to increased fracture risk and reduced quality of life. While traditional risk factors such as age, hormonal changes, and lifestyle are well established, the impacts of diabetes and obesity on osteoporosis remain unclear. This study aimed to investigate associations between diabetes, obesity, and osteoporosis diagnosis in Caucasian women aged 64 years and older.

**Materials and Methods:** Data on osteoporosis diagnosis, diabetes diagnosis, and body mass index (BMI) were obtained from the publicly available Study of Osteoporotic Fractures (SOF) database. Statistical analyses were conducted using IBM SPSS software. Associations between diabetes, BMI, and osteoporosis were evaluated at two study visits (visit 1 and visit 8). Analysis of variance (ANOVA) and correlation analyses were used to assess relationships among variables.

**Results:** No significant association was found between diabetes and osteoporosis at visit 1 (p = 0.966); however, a statistically significant association emerged at visit 8 (p < 0.001). A weak negative correlation between diabetes and osteoporosis was observed at visit 8 (r = –0.068, p < 0.001), indicating that participants with diabetes were slightly less likely to be diagnosed with osteoporosis. BMI category was significantly associated with osteoporosis at both visits (p < 0.001). Post hoc analyses revealed that overweight and obese women had a lower likelihood of osteoporosis than underweight or normal-weight participants.

**Conclusions:** Diabetes showed no consistent association with osteoporosis diagnosis, whereas higher BMI appeared to exert a protective effect against osteoporosis in older women.

## 1 INTRODUCTION

Osteoporosis is a common yet burdensome disease that affects both men and women, particularly in older adulthood. It is estimated that 1 in 2 women and 1 in 4 men over the age of 50 will experience an osteoporosis-related fracture in their lifetime [1]. In the United States alone, approximately 10.2 million adults are diagnosed with osteoporosis, with women accounting for 80% of those cases [2, 3]. A 2021 systematic review and meta-analysis examining data from 40 studies across Asia, Europe, and the Americas found that 21.7% of older adults globally are affected by osteoporosis. Prevalence rates were 35.3% in women and 12.5% in men, with the highest rates observed in Asia at 24.3% [4]. In the United States, recent national data indicates that Asian non-Hispanic adults over 50 have the highest prevalence at 18.4%, followed by non-Hispanic White adults at 12.9%, and non-Hispanic Black adults at 6.8% [5]. While prior studies have identified numerous contributing factors to osteoporosis diagnosis, ongoing research continues to explore additional associations to enable earlier detection and improve long-term health outcomes and quality of life for individuals with osteoporosis.

Osteoporosis is widely recognized as a “bone weakening disease” however, the disease is much more complicated as there are many intricate changes that occur leading to its development. As a process of keeping bones healthy, osteoclasts, a type of cell, break down old bone while another type of cell, osteoblasts, form new bone. The normal structure inside of a bone appears like a honeycomb, with noticeable bone forming the walls of the honeycomb. However, the bone forming the walls in an individual with osteoporosis becomes much smaller and the spaces between bone grow larger. The outer shell of the bone also becomes thinner, thus making the bone significantly weaker [6].

“Silent Diseases” are diseases which often remain asymptomatic until severe symptoms develop. Osteoporosis falls into this category of diseases as individuals often do not have symptoms until a fracture occurs or the disease has significantly progressed [2]. Symptoms of the disease can include frequent bone fractures, fractures which occur with minimal force, back pain, loss of height, and bone pain – most commonly in the spine [7]. Osteoporotic fractures, a common symptom, significantly contribute to increased morbidity and mortality, while also leading to a marked decline in patients’ overall quality of life [8].

Treatment options include both lifestyle modifications and medications. Regarding lifestyle changes, physicians may recommend increasing one’s exercise, especially weight-bearing activities and those that improve balance. Improving balance and increasing overall fitness, often decreases the likelihood of falling and fracturing a bone. Other lifestyle changes include the cessation of smoking, limiting alcohol intake, and consuming enough calcium and vitamin D through proper nutrition or supplements [7,9]. Research has demonstrated the importance of adequate nutrition, particularly consuming enough vitamin D and calcium, in preventing and treating osteoporosis [1].

Since the 1970s, various medications have been developed to treat osteoporosis in both men and women. These medications include antiresorptive or bisphosphonate drugs, and anabolic or bone building drugs. Current and common antiresorptive drugs on the market include; Alendronate, Risendronate, Ibandronate, Zoledronic acid, and Denosumab. Current anabolic medications include; Teriparatide, Abaloparatide, and Romosozumab [9]. Additionally, research has shown estrogen-replacement therapy to be a successful treatment option for postmenopausal women [10]. This treatment option is particularly important considering osteoporosis is estimated to affect 200 million women across the world [11]. There are different criteria that clinicians take into consideration when determining which medication is best for a patient. Some criteria may include the patient’s biological sex, DXA scan results, age of the patient, comorbidities, degree of fracture risk, and other medication use. Different osteoporosis medications have shown to help prevent different types of fractures, which may be considered when looking into a patient’s history and future risk of fractures. Lastly, cost-effectiveness is often considered, with oral bisphosphonates, excluding ibandronate, often being prescribed as a first-line treatment option [12,13].

Diabetes mellitus and obesity are two major factors implicated in the development of many diseases. Diabetes results from inadequate insulin production or ineffective insulin utilization, depending on the type of disease. Type 1 is characterized as an autoimmune disease where pancreatic beta cells are completely destroyed. Type 1 accounts for between 5-10% of cases of diabetes. Type 2 is characterized by insulin resistance where insulin is ineffective. Type 2 accounts for 90% of cases of diabetes. Insulin, a peptide hormone, plays several essential roles in the body, most notably, regulating blood glucose levels [14]. A systematic review and meta-analysis from 2023 looked into the prevalence of osteoporosis in individuals with type 2 diabetes specifically and found that 27.67% (95% CI 21.37 – 33.98%) of those with diabetes had osteoporosis [15]. With that data in mind and also given that 11.6% of the United States population is living with diabetes, further research is essential to understand the disease and its associated risks [16].

Obesity is determined by a measurement that takes into account an individual’s weight and height, otherwise known as bone mineral index (BMI). The BMI measurement was first established in the 19^th^ century, but has been re-defined since 1972. Over the years, there has been much controversary about using BMI as the standard for determining obesity. Researchers have suggested that a waist-to-hip ratio (WHR) is more accurate in determining the health and risk of illness in individuals [17]. Regardless, BMI charts currently define if one is overweight (25-29.9) or obese (>30) [18]. Approximately 31% of adults in the U.S. population are overweight, over 42% are obese, and 9.2% classified as morbidly obese [19]. Given the rising obesity rates over the past two decades, its health effects have become a critical focus of study.

Several factors contribute to the development of osteoporosis, including age, diet, family history, medication use, hormonal changes, body size, and lifestyle [7]. This study investigates how diabetes and obesity may influence the risk of osteoporosis in Caucasian women over the age of 64. By analyzing these variables separately, the study aims to determine whether a diagnosis of diabetes or obesity is associated with an increased or decreased likelihood of osteoporosis. Understanding these relationships may help guide more targeted screening and prevention strategies for aging populations.

## 2 MATERIALS AND METHODS

### 2.1 |Data Source and Study Population

All data for this analysis was collected from the Study of Osteoporotic Fractures (SOF) online database, which contains data from a multicenter observational study of 9,704 women. In 1986, the National Institutes of Health began the SOF research study by collecting demographic information, health histories, and biospecimens from consenting women over the age of 64. All women were Caucasian in the study until 1997, the sixth visit, when an additional 662 African American women were enrolled [20]. All data collected for this study was from Caucasian women. The details of inclusion/exclusion criteria for the studies are described in Figure 1, following the Preferred Reporting Items for Systematic Reviews and Meta-Analyses (PRISMA) guidelines.

**FIGURE 1.**
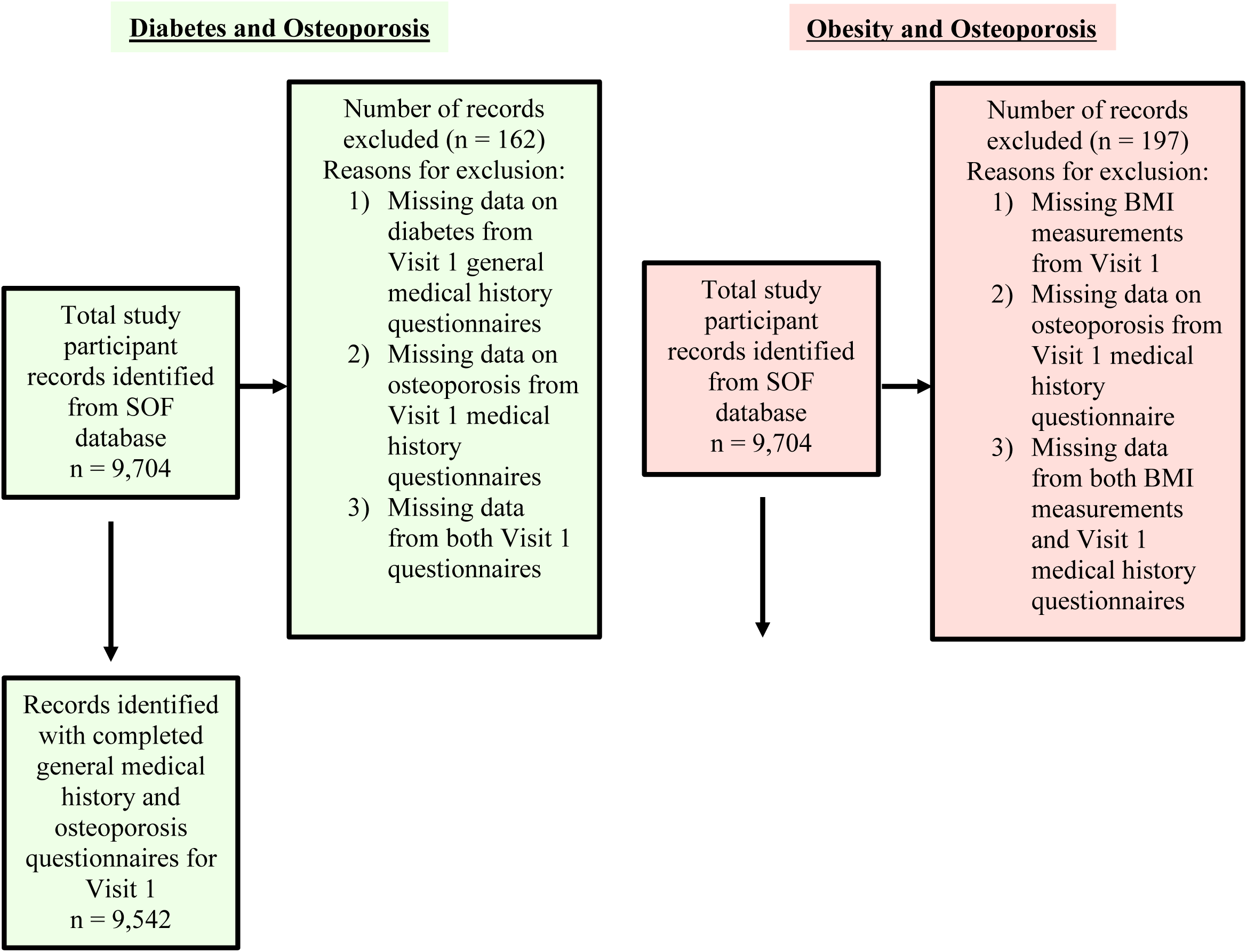
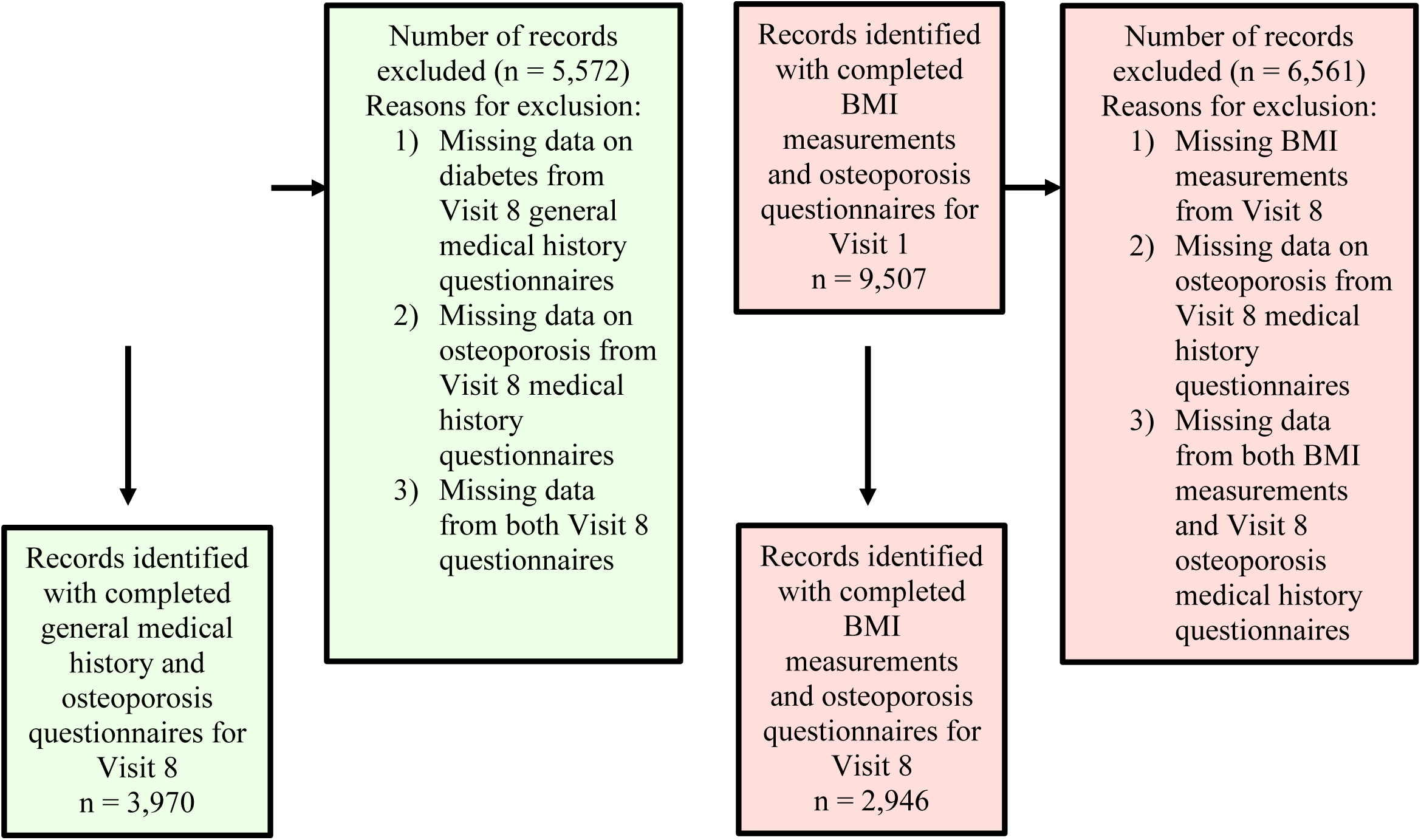
PRISMA Methods for Collecting Data on Diabetes and Osteoporosis and Obesity and Osteoporosis at Visits 1 and 8.

### 2.2 Standardized questionnaires

Participants were contacted for follow-up every two years. At each visit, anthropometric measurements were collected, and participants completed standardized questionnaires regarding new health diagnoses, including diabetes and osteoporosis. For this analysis, data from visit 1 and visit 8 were selected. Self-reported diagnoses of diabetes and osteoporosis were obtained from the questionnaires administered at both visits. Height and weight were measured by study teams, and BMI was calculated [20]. Figures S1 through S4 in the supplemental information document detail the specific questionnaire items administered to participants as part of the data collection process for this study.

### 2.3 BMI Categories

BMI categories for this analysis were determined based on the Centers for Diseases Control and Prevention (CDC) BMI guidelines as shown in Table S1 [18]. All raw data for participants BMI was categorized into values 0-5 for each of the BMI categories: 0 = underweight, 1 = Healthy Weight, 2 = Overweight, 3 = Class 1 Obesity, 4 = Class 2 Obesity, and 5 = Class 3 Obesity.

### 2.4 Statistical Analysis

Data was downloaded from the SOF online database and converted from SAS file format to Microsoft Excel for compatibility with IBM SPSS Statistics software. All statistical analyses were conducted using IBM SPSS Software.

## 3 RESULTS

Table 1 presents age-related demographic data for participants in the SOF study at visit 1. The majority of women enrolled were between 65–69 years old (n = 4,113), followed by those aged 70–74 (n = 3,044). Only 200 participants were in the 85–89 age group at the time of the first study visit. This table 1 provides a general overview of the age distribution within the study population.

**TABLE 1.** Participant Age Demographics, Duration Since Diabetes Diagnosis, and Number of Osteoporosis Diagnoses Among Women with Diabetes at Visit 1.

| **Demographic Data: All Study Participants** |  |
| --- | --- |
| **Age Group** | **Number of Participants** |
| 65-69 years | 4,113 |
| 70-74 years | 3,044 |
| 75-79 years | 1,543 |
| 80-84 years | 773 |
| 85-89 years | 200 |
| No Answer | 31 |
| Total | 9,704 |
| **Diabetes Diagnoses: Visit 1** |  |
| **Years Since Diabetes Diagnosis** | **Number of Participants** |
| 0-10 years | 413 |
| 11-20 years | 170 |
| 21-30 years | 68 |
| 31-40 years | 19 |
| 41+ years | 10 |
| All Years | 680 |
| **Osteoporosis Diagnoses in Women with Diabetes: Visit 1** |  |
| **Years Since Diabetes Diagnosis** | **Number of Participants also with Osteoporosis Diagnoses** |
| 0-10 years | 53 |
| 11-20 years | 30 |
| 21-30 years | 13 |
| 31-40 years | 2 |
| 41+ years | 1 |
| All Years | 99 |

Additionally, Table 1 includes data on the 680 participants who had been diagnosed with diabetes at visit 1. Most of these individuals (n = 413) had received a diabetes diagnosis within the past 0–10 years, followed by 170 participants diagnosed 11–20 years prior. While the diabetes diagnosis was not specified by type (type 1 or type 2), the age of study participants (all over 64 years) suggests that those with a diagnosis more than 41 years prior may have had type 1 diabetes.

Lastly, Table 1 details the subset of 99 participants who had been diagnosed with both diabetes and osteoporosis at visit 1. Among these, 53 had been diagnosed with diabetes within the previous 10 years, and 30 had received a diagnosis 11–20 years before visit 1.

A chi-squared test of independence was conducted to evaluate the relationship between participant diabetes diagnosis and osteoporosis diagnosis at both visit 1 and visit 8 in Table 2. At visit 1, the results showed no statistically significant association between diabetes and osteoporosis diagnoses at visit 1, χ²(1, N = 9,542) = 0.002, *p* = 0.966. However, a significant association was found at visit 8, χ²(1, N = 3,970) = 18.094, *p* < 0.001, indicating a potential relationship between the two variables at that visit. Additionally, Pearson’s correlation analysis revealed a very weak negative correlation between diabetes and osteoporosis diagnoses (*r* = –0.068, *p* < 0.001), suggesting that individuals with diabetes were slightly less likely to be diagnosed with osteoporosis. Table S2 in the supplemental information document displays the symmetric measures from Pearson’s correlation analysis.

**TABLE 2.** Crosstabulation Tables for Osteoporosis Diagnosis and Diabetes Diagnosis at Visits 1 and 8 and Pearson Chi-Squared Results.

| **<u>Visit 1</u>** |  |  |  |  |
| --- | --- | --- | --- | --- |
|  |  | **No Osteoporosis**<br>**Diagnosis** | **Osteoporosis**<br>**Diagnosis** | **Total** |
| **No Diabetes Diagnosis** |  | 7,562 | 1,314 | 8,876 |
| **Diagnosed with**<br>**Diabetes** |  | 567 | 99 | 666 |
| **Total** |  | 8,129 | 1,413 | 9,542 |
|  |  | **Value** | **df** | **Asymptotic**<br>**Significance (2-sided)** |
| **Pearson Chi-Squared** |  | 0.002 | 1 | 0.966 |
| **Continuity Correction** |  | 0 | 1 | 1 |
| **Likelihood Ratio** |  | 0.002 | 1 | 0.966 |
| **Linear-by-Linear**<br>**Association** |  | 0.002 | 1 | 0.966 |
| **N of Valid Cases** |  | 9,542 |  |  |

| **<u>Visit 8</u>** |  |  |  |  |
| --- | --- | --- | --- | --- |
|  |  | **No Osteoporosis**<br>**Diagnosis** | **Osteoporosis**<br>**Diagnosis** | **Total** |
| **No Diabetes Diagnosis** |  | 2,373 | 1,207 | 3,580 |
| **Diagnosed with**<br>**Diabetes** |  | 300 | 90 | 390 |
| **Total** |  | 2,673 | 1,297 | 3,970 |
|  |  | **Value** | **df** | **Asymptotic**<br>**Significance (2-sided)** |
| **Pearson Chi-Squared** |  | 18.094 | 1 | <0.001* |
| **Continuity Correction** |  | 17.614 | 1 | <0.001 |
| **Likelihood Ratio** |  | 19.116 | 1 | <0.001 |
| **Linear-by-Linear**<br>**Association** |  | 18.089 | 1 | <0.001 |
| **N of Valid Cases** |  | 3,970 |  |  |

### Obesity and Osteoporosis

Figure 2 displays the participants BMI categories as a percentage for visits 1 and 8, due to the study populations being different in number. The majority of participants at visit 1 had a BMI classified as “healthy” (41.73%) while the majority of participants at visit 8 had a BMI classified as “overweight” (38.8%). In the supplemental information document, figures S5 and S6 provide raw data for BMI categorization at visits 1 and 8.

**FIGURE 2.**
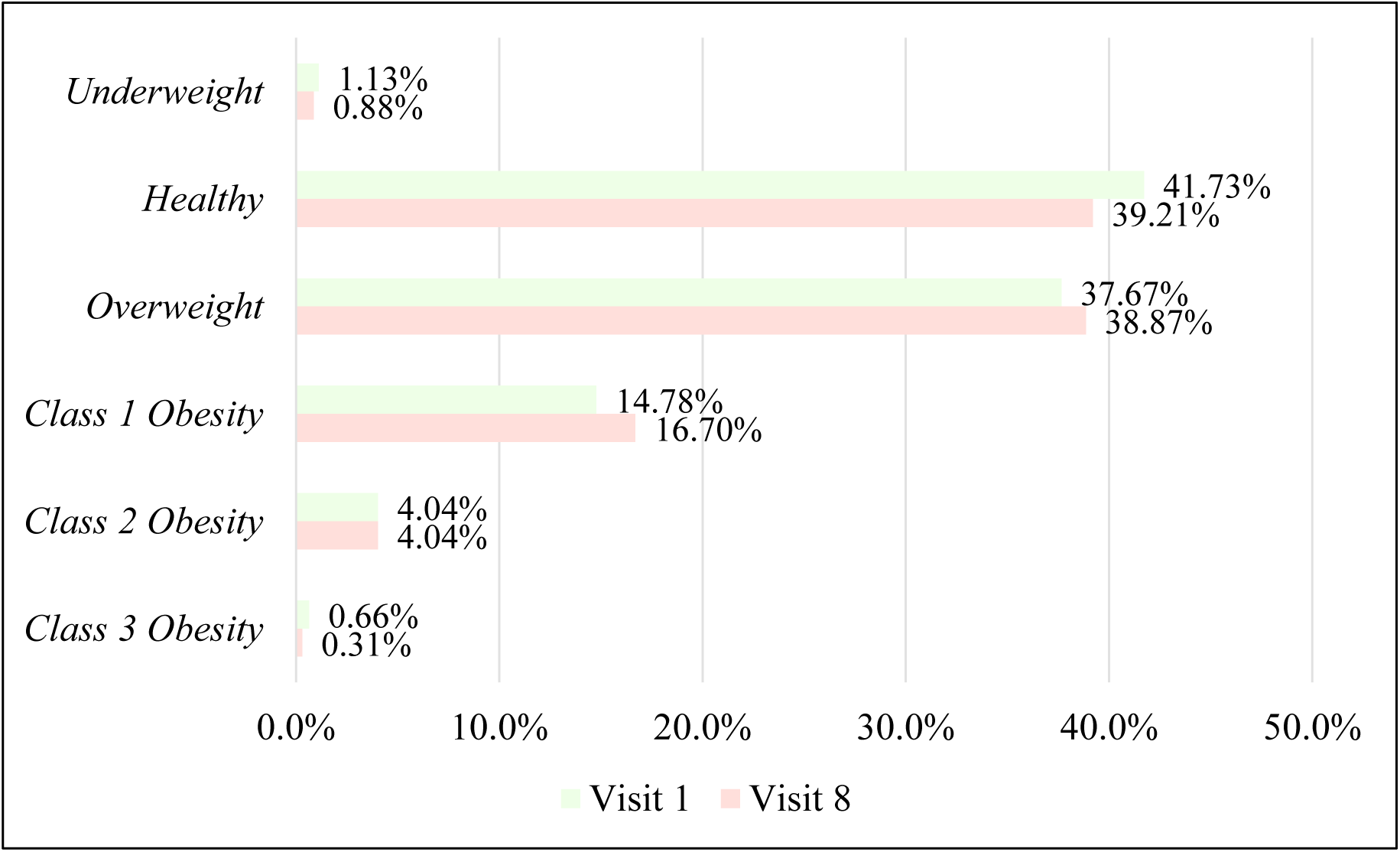
Percentage of Participants in Each BMI Category at Visits 1 and 8.

A chi-squared test of independence was conducted to evaluate the relationship between participant BMI category and osteoporosis diagnosis at both visit 1 and visit 8 in Table 3. At visit 1, the relationship was statistically significant, χ²(5, N = 9,507) = 21.533, *p* < 0.001. A one-way ANOVA test also revealed a significant difference in osteoporosis diagnosis across BMI categories (F = 4.314, *p* < 0.001), with post-hoc analysis showing significant differences between underweight and class 2 obesity (*p* = 0.044), and underweight and class 3 obesity (*p* = 0.049). At visit 8, the chi-squared test again indicated a statistically significant association between BMI category and osteoporosis diagnosis, χ²(5, N = 2,946) = 43.810, *p* < 0.001. The one-way ANOVA test further supported this, showing a significant difference across BMI categories (F = 8.876, *p* < 0.001). Post-hoc comparisons revealed significant differences between healthy weight and overweight (*p* = 0.004), healthy weight and class 1 obesity (*p* < 0.001), and healthy weight and class 2 obesity (*p* < 0.001). In the supplemental information document, tables S3 and S4 provide all post-hoc ANOVA results with significant values found from visit 1 and 8 data analysis.

**TABLE 3.** Crosstabulation Tables for Osteoporosis Diagnosis and BMI Category at Visits 1 and 8 and Pearson Chi-Squared Results.

| **<u>Visit 1</u>** |  |  |  |
| --- | --- | --- | --- |
|  | **No Osteoporosis**<br>**Diagnosis** | **Osteoporosis**<br>**Diagnosis** | **Total** |
| **Underweight** | 83 | 24 | 107 |
| **Healthy Weight** | 3,330 | 637 | 3,967 |
| **Overweight** | 3,067 | 514 | 3,581 |
| **Class 1 Obesity** | 1,222 | 183 | 1,405 |
| **Class 2 Obesity** | 341 | 43 | 384 |
| **Class 3 Obesity** | 59 | 4 | 63 |
| **Total** | 8,102 | 1,405 | 9,507 |
|  | **Value** | **df** | **Asymptotic**<br>**Significance (2-sided)** |
| **Pearson Chi-Square** | 21.533 | 5 | <0.001* |
| **Likelihood Ratio** | 22.092 | 5 | <0.001 |
| **Linear-by-Linear**<br>**Association** | 19.202 | 1 | <0.001 |
| **N of Valid Cases** | 9,507 |  |  |

| **<u>Visit 8</u>** |  |  |  |
| --- | --- | --- | --- |
|  | **No Osteoporosis**<br>**Diagnosis** | **Osteoporosis**<br>**Diagnosis** | **Total** |
| **Underweight** | 15 | 11 | 26 |
| **Healthy Weight** | 733 | 422 | 1,155 |
| **Overweight** | 806 | 339 | 1,145 |
| **Class 1 Obesity** | 374 | 118 | 492 |
| **Class 2 Obesity** | 100 | 19 | 119 |
| **Class 3 Obesity** | 5 | 4 | 9 |
| **Total** | 2,033 | 2,946 | 2,946 |
|  | **Value** | **df** | **Asymptotic Significance (2-sided)** |
| **Pearson Chi-Square** | 43.81 | 5 | <0.001* |
| **Likelihood Ratio** | 45.391 | 5 | <0.001 |
| **Linear-by-Linear Association** | 38.916 | 1 | <0.001 |
| **N of Valid Cases** | 2,946 |  |  |

## 4 DISCUSSION

The relationship between diabetes and osteoporosis yielded both statistically significant and non-significant findings across study visits. At visit 1, no association was observed between diabetes diagnosis and osteoporosis. However, by visit 8, an inverse correlation emerged, suggesting that women with diabetes may have a slightly reduced risk of developing osteoporosis. While this inverse relationship reached statistical significance, the effect size was minimal and may lack clinical relevance without further investigation. These findings align with existing literature, which has identified diabetes, particularly type 2, as both a potential risk factor and, paradoxically, a possible protective factor against osteoporosis in certain contexts. [21,22]. Additionally, a 2007 study found that the odds of developing osteoporosis were 22.54% higher for those with type 2 diabetes compared to those without diabetes [23].

A 2021 systematic review and meta-analysis including 24,340 patients, revealed no relationship between type 1 and type 2 diabetes and low bone density [24]. This is important in relation to our study as low bone density is correlated with the development of osteoporosis. Therefore, the results of our study align with the 2021 meta-analysis. In another large-scale retrospective cohort involving Taiwanese adults diagnosed with type 2 diabetes between 2002 and 2015, researchers found that patients with type 2 diabetes had a significantly elevated risk of osteoporosis, with a hazard ratio of 1.37 (95% CI: 1.11–1.69) [25]. A recent meta-analysis by Cao et al. (2025) also reported that individuals with type 2 diabetes have a higher risk of osteoporosis (RR = 1.841; 95% CI: 1.219–2.780; *p* = .004), along with a greater fracture risk (RR = 1.21; 95% CI: 1.09–1.31; *p* < .001) compared to non-diabetic individuals [26].

The association between obesity and osteoporosis, however, was consistently significant. Overweight and obese women had lower odds of developing osteoporosis—a finding supported by prior meta-analyses. A 2023 systematic review found that overweight and obese individuals had reduced odds of osteoporosis (OR = 0.451, CI = 0.366–0.557), while underweight individuals had significantly increased risk (OR = 2.540, CI = 1.483–4.350) [27].

Studies dating back to 2006 even support our research findings. Asomaning et al. (2006) conducted a cross-sectional study of postmenopausal women aged 50–84 and found a clear inverse relationship between BMI and osteoporosis. Specifically, compared to women with moderate BMI, those categorized as low BMI had 1.8 times higher odds of osteoporosis (OR = 1.8; 95% CI: 1.2–2.7), while overweight women had significantly reduced odds (OR = 0.46; 95% CI: 0.29–0.71), and obese women had even lower odds (OR = 0.22; 95% CI: 0.14–0.36). In continuous analysis, each one-unit increase in BMI corresponded to a 12% reduction in osteoporosis risk (OR = 0.88; 95% CI: 0.85–0.91). These results emphasize that lower BMI is associated with elevated osteoporosis risk, while higher BMI confers a protective effect independent of major lifestyle or hormonal factors. [28].

A second cross-sectional study using data from 10,942 Taiwanese adults, Wu et al. (2022) found a strong association between body mass index (BMI) and osteoporosis risk. Compared to individuals with a normal BMI, those who were underweight (BMI < 18.5 kg/m²) had significantly increased odds of developing osteoporosis (OR = 6.52; 95% CI: 4.62–9.19). Conversely, overweight individuals (BMI 24–27 kg/m²) showed substantially decreased odds (OR = 0.176; 95% CI: 0.140–0.221), and those who were obese (BMI ≥ 27 kg/m²) had the lowest risk, with an odds ratio of 0.057 (95% CI: 0.039–0.083). These findings clearly suggest a protective effect of higher BMI against osteoporosis, while underweight status dramatically increases one’s risk [29].

Although our study produced many significant findings, there are limitations that must be discussed. Limitations of this study include sample size attrition by visit 8, likely due to mortality or study withdrawal. The smaller sample size by visit 8 may have had an effect on results. Additionally, many of the questions on participant questionnaires were self-reported and recall bias could have been introduced.

Another limitation was the lack of distinction in the datasets between type 1 and type 2 diabetes. The two forms differ significantly in etiology and typical age of onset, which may influence osteoporosis risk differently. Literature also indicates a significant difference in bone deteriorations between type 1 and type 2 diabetes due to differing molecular mechanisms [30]. This is worth noting, as differing bone deterioration patterns have potential to influence osteoporosis development differently.

Only Caucasian women were analyzed in this study, as African American participant data were kept separate after their enrollment at visit 6. Introducing other racial groups in future analyses may yield different results. Lastly, only women were included in the SOF study, so including men could give different results on the associations with our variables and osteoporosis.

It is evident that there are significant associations between osteoporosis and obesity, with obesity being a protective effect. This may seem contradictory to research on the negative impacts of obesity on one’s health. However, as mentioned, previous literature and reported studies support the analyses in our study. There are several mechanisms which infer as to why obesity can be considered a protective affect against osteoporosis.

In summary, first, overweight and obese women have increased estrogen synthesis. Estrogen, a steroid hormone, is very important in bone health and helps to reduce the breakdown of bone while also stimulating bone formation [31]. This is why estrogen-replacement therapy in postmenopausal women can be beneficial. By having increased fat tissue and thus higher amounts of estrogen, the risk of osteoporosis development should be lowered. Secondly, research has demonstrated that total body fat is positively related to bone mineral density. Thus, women with a higher BMI will have higher bone mineral densities, thus decreasing the risk of developing osteoporosis. Lastly, women with higher BMI’s have an increased mechanical load and strain. This ultimately strengthens the bone and helps to prevent fractures and the deterioration of bone [32]. Maintaining a healthy BMI throughout one’s life remains important and overall has many positive benefits on one’s health. However, clinicians should recognize that overweight and obese women have a decreased odds of developing osteoporosis, and they should council patients, especially postmenopausal women, appropriately.

## Data Availability

The data associated with this paper is publicly available at https://sofonline.ucsf.edu/.

## AUTHOR CONTRIBUTIONS

**Mendon G. Thomas** contributed to data curation, data analysis and interpretation, and initial manuscript drafting. **Ambalangodage C. Jayasuriya** conceived and designed the study, supervised the project, and reviewed and edited the manuscript. All authors contributed to the article and approved the final version of the manuscript.

## ACKNOWLEDGEMENTS

The authors declare no acknowledgement.

## DATA AVAI LAB ILITY STATEMENT

The data associated with this paper is publicly available at https://sofonline.ucsf.edu/.

## ETHICS STATEMENT

This study used a SOF database and did not involve any human participants, animal subjects, or patient data.

